# Exploring scalable assessment methods for terminated trials in ClinicalTrials.gov: A cohort analysis of German and Californian trials

**DOI:** 10.1101/2025.07.08.25330734

**Authors:** Samruddhi Suresh Yerunkar, Benjamin Gregory Carlisle, Delwen L. Franzen, Maia Salholz-Hillel, Daniel Strech, Susanne Gabriele Schorr

## Abstract

**Introduction:** Clinical trials can terminate early for many reasons, including non-scientific reasons. We aimed to develop scalable semi-automated methods to characterize terminated trials and explore a methodology to estimate the risk of experiencing serious adverse events (SAEs) in trials terminated due to non-scientific reasons.

**Methods:** Two cohorts of clinical trials registered in ClinicalTrials.gov were investigated: (1) a cohort of clinical trials affiliated with German university medical centres (reported as completed between 2009-2017) and (2) a cohort of clinical trials affiliated with Californian university medical centers (reported as completed between 2014-2017). We used these cohorts to explore scalable assessment methods and compare terminated trials to completed ones regarding trial characteristics, including therapeutic focus. In a subset of trials terminated for non-scientific reasons with tabular summary results and a parallel, randomized design, we estimated additional risk for SAE. For the German cohort, if results were missing from ClinicalTrials.gov, results from the EU Clinical Trials Register (EUCTR) were included if available.

**Results:** Of 2,253 German and 1,091 Californian trials, 217 (10%) and 150 (14%) were terminated, respectively. The majority (German: 65%, Californian: 67%) cited non-scientific reasons for termination, primarily low accrual. Compared to completed trials, terminated trials showed lower rates of results reporting: 35% vs. 78% in Germany and 72% vs. 87% in California. Of 242 trials terminated for non-scientific reasons, 14 (6%; 11 from ClinicalTrials.gov, 3 from EUCTR) could be included in the SAE risk assessment. No significant difference in SAE risk was observed between the intervention and control arms (RR 1.05, 95% CI: 0.76-1.44).

**Discussion:** Results of terminated trials were less frequently reported, limiting opportunities for knowledge generation. Broader adoption of harmonized result reporting standards across registries, structured templates, and improved logic checks could enable scalable assessment approaches and enhance the utility of terminated trial data for clinical research transparency.

## Introduction

Clinical trials are the cornerstone of evidence-based healthcare, providing essential information on the safety and efficacy of new treatments and interventions. Rigorously planned and conducted randomized clinical trials are critical for informing healthcare decisions (1), with various guidelines in place to ensure they are conducted with the highest standards of safety, ethics, and integrity. Besides informing health care decisions, results and methods used in clinical trials are also relevant in shaping the design of new studies (2, 3). However, despite their importance and structured planning processes, not all clinical trials can reach their pre-defined goals, and some discontinue prematurely (terminate).

Speich et al. (2022) analyzed 326 trials approved by research ethics committees in Switzerland, the United Kingdom (UK), Germany, and Canada and reported that 30% (98/326) of trials were prematurely discontinued. The study also highlights that prematurely discontinued trials were less likely to have their results published in journals or reported in registries compared to completed trials (1). Premature discontinuation of trials can occur due to scientific reasons, such as ethical concerns related to efficacy or safety, or non-scientific reasons such as poor project planning or low enrollment rates (4, 5). Williams et al. (2015) highlighted that, as of February 2013, the most common reason for trial termination among terminated trials was non-scientific reasons (5). This is particularly concerning because this might, in principle, have been foreseen or prevented, leading to unnecessary expenditure of time and resources. Most importantly, participants who invest their time to participate in clinical research might be exposed to interventions and measures with unclear benefits, and risk experiencing adverse events.

Clinical trial interventions are often experimental and carry a higher risk of adverse events than standard clinical practice (6). Serious adverse events (SAEs) are defined by ClinicalTrials.gov as ‘any untoward medical occurrence that at any dose results in death, is life-threatening, requires inpatient hospitalization or prolongation of existing hospitalization, results in persistent or significant disability/incapacity, or is a congenital anomaly/birth defect’ (7). Studies have consistently shown that reporting of SAEs is more comprehensive on ClinicalTrials.gov compared to reporting of SAEs in published journal articles (8–10), enabling better safety data synthesis. Assessing SAE data available in such trial registries can offer additional context for understanding safety aspects in clinical research (11). Given that most premature terminations occur for non-scientific reasons, investigating SAE data from these trials may help to understand the potentially avoidable risk faced by participants enrolled in these trials.

Our group has previously investigated how University Medical Centers (UMC) affiliated clinical trials in Germany and California perform on transparency practices, such as prospective registration and results reporting (3, 12, 13). For prematurely discontinued trials in those cohorts, we did not assess the reason for termination or how they differed from completed trials. In this study, we aimed to develop scalable semi-automated methods to characterize terminated trials in these two cohorts, quantify reasons for termination, and explore a methodology to estimate the risk of experiencing SAEs in trials terminated due to non-scientific reasons.

## Materials and Methods

The protocol for this project was preregistered on the Open Science Framework (OSF) on November 21, 2023, and is available at https://osf.io/n4ujs/. The study is reported according to the Strengthening the Reporting of Observational Studies in Epidemiology (STROBE) guideline for cross-sectional studies (14).

### Data sources

We used two cohorts of clinical trials: (a) those affiliated with German UMCs and (b) those affiliated with Californian UMCs. The German cohort includes interventional clinical trials registered on ClinicalTrials.gov or the German Clinical Trials Register (Deutsches Register Klinischer Studien, DRKS), which were affiliated with a German UMC and reported as complete between 2009 and 2017. This dataset is openly available on GitHub (https://github.com/maia-sh/intovalue-data) (15). The Californian cohort includes interventional clinical trials registered on ClinicalTrials.gov, affiliated with a Californian UMC and reported as complete between 2014 and 2017, and is openly available on GitHub (https://github.com/ontogenerator/california-clinical-dashboard) (16). For both cohorts, clinical trial results publications have been searched and validated manually. These datasets were selected due to their public availability and prior manual results publication searches.

### Eligibility criteria

We included all trials from these datasets that were registered on ClinicalTrials.gov, had a recruitment status ‘terminated’ as of the historical version downloaded on December 1, 2023, using the cthist R package (17), and at least one participant enrolled. ClinicalTrials.gov defines ‘terminated’ as trials where the recruitment or enrollment of participants has halted prematurely and will not resume, and participants are no longer being examined or receiving intervention. We compared the characteristics of these terminated trials with those of trials from the same cohort with a recruitment status of ‘completed’. ClinicalTrials.gov defines ‘completed’ trials as those in which the study concluded as planned, and participants are no longer receiving intervention or being examined (i.e., the last participant’s final visit has occurred) (18).

### Data extraction and analyses

Characteristics of the trials (phase, manually identified results publications) were taken from the original datasets. To generate version-specific variables for assessing terminated trials, we developed a <u>custom R package named “terminated_trials_study”</u> which is openly available on GitHub under the GNU Affero General Public License v3.0 (AGPL-3.0). This package uses the <u>cthist R package</u> (17) to download clinical trial registry entry histories in a structured format. Based on this data, we generated the following variables: degree of enrollment, trial days, summary results and reason for termination. In addition, we developed a novel R package, TrialFociMapper (19), to assign therapeutic foci for trials registered in ClinicalTrials.gov.

‘Degree of enrollment’ was defined by the ratio of actual and anticipated enrollment, expressed as a percentage of enrollment when the trial is terminated. In ClinicalTrials.gov, the enrollment variable is ‘estimated’ at the start of the trial and can be updated to ‘actual’ at various stages of the trial, including at its completion/termination. The function defines anticipated enrollment as the estimated enrollment reported in the first version on or after the trial’s start date and actual enrollment as actual enrollment recorded at the end of the trial. If either anticipated or actual was missing, the function flagged a warning about the missing data. For records where an anticipated number was specified but not explicitly marked as anticipated, we manually assigned it as anticipated. Other records with unclear or missing enrollment information were excluded. <u>See appendix 1 for details</u>.

‘Trial days’ were defined by the number of days between the trial’s stop and start dates. The stop date is defined as the primary completion date reported when its overall status was first changed to terminated from any other status. The ‘start date’ is obtained from the same version of the trial record as the stop date.

The availability of summary results was assessed using the final version of the trial record on ClinicalTrials.gov. Trials investigating investigational medicinal products in Germany are legally required to be registered and to report summary results in the EU Clinical Trials Register (EUCTR). For this German cohort, we additionally applied a custom scraper function to extract summary results from EUCTR. This allowed us to capture additional reporting that may not have been available on ClinicalTrials.gov. Summary results were considered available if present in either ClinicalTrials.gov or the EUCTR registry.

We automatically retrieved the reasons for termination from the ClinicalTrials.gov registry entry ‘why_stopped’ field. This field contains free-text explanations (limited to 160 characters). Two authors (SGS and SSY) manually categorized reasons for termination using a categorization table developed based on existing studies and external sources (1, 4, 5, 20–23). If both non-scientific and scientific reasons were mentioned, the scientific reason was considered primary. In cases where multiple scientific or multiple non-scientific reasons were provided, the description and sequence of reasons were used as a reference (<u>see appendix</u> table 1<u>, Reason for termination categorization</u>). Discrepancies between authors were resolved by discussion, and if no agreement was reached, a third author (BGC) was consulted. Inter-rater agreement was calculated for the agreement between the two raters (SGS and SSY).

**Table 1:**
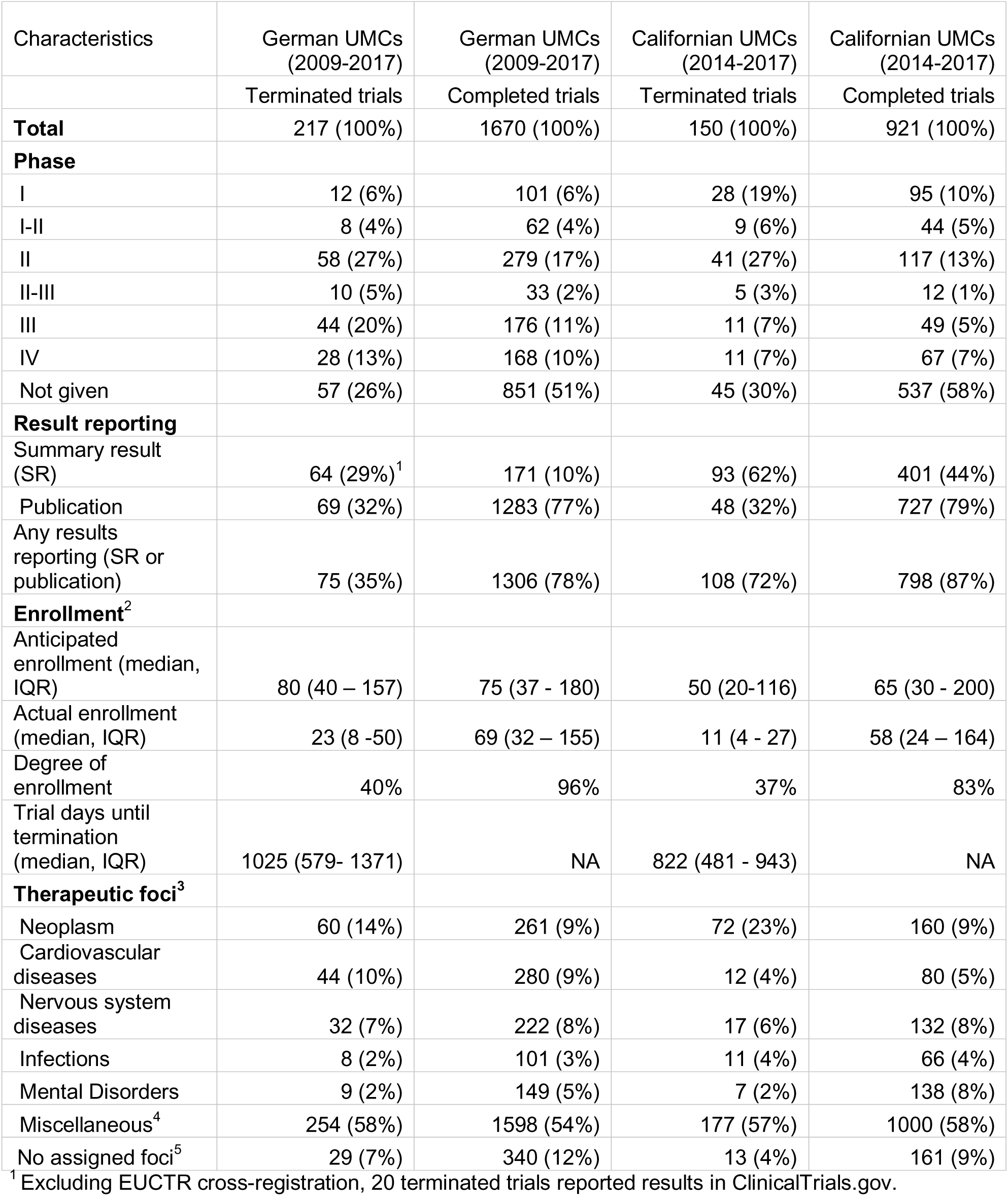
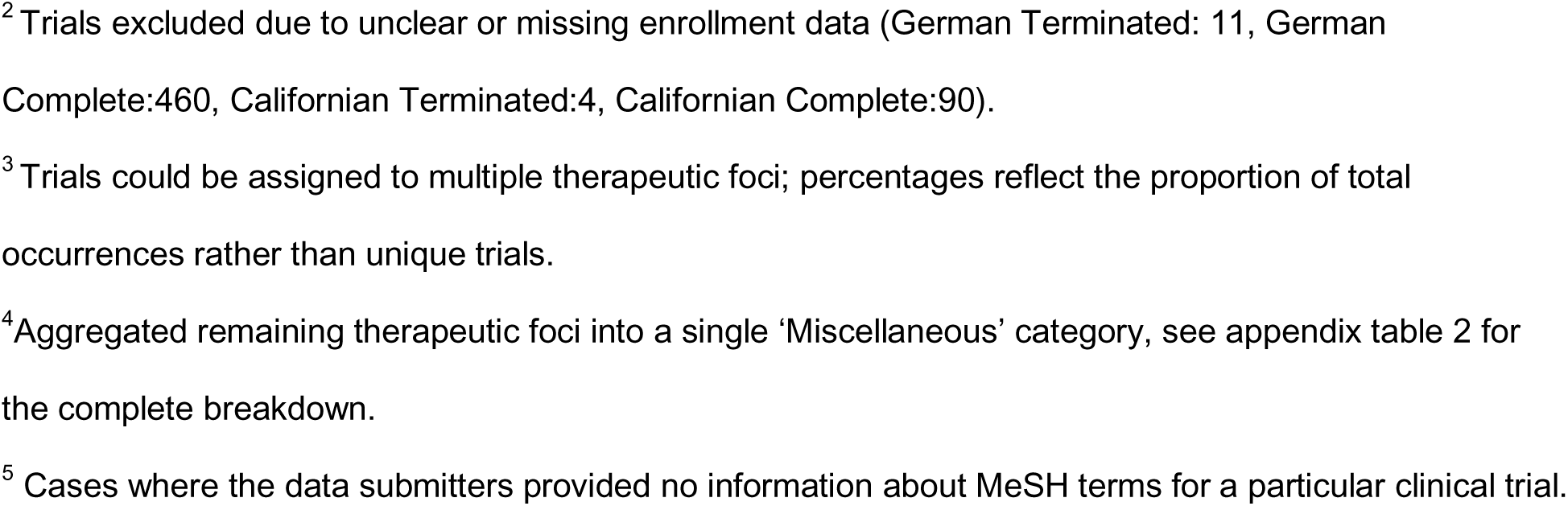
Characteristics of terminated trials and completed trials.

The TrialFociMapper package retrieves the ‘browse_conditions’ table from the AACT (Aggregate Analysis of ClinicalTrials.gov) (24) database, which contains Medical Subject Headings (MeSH) terms submitted by data submitters. These MeSH terms, developed by the National Library of Medicine (NLM), are mapped to high-level categories based on the hierarchy of the MeSH tree. For instance, if a data submitter provides “breast cyst” as a MeSH term, the function assesses the MeSH tree to map “breast cyst” to “neoplasms”, under which it is categorized in the MeSH tree. A trial can have no focus (if not submitted by the data submitter), one focus, or multiple foci. We reported how often each of the therapeutic foci appears across all trials, allowing for multiple foci per study.

### Serious adverse events (SAE)

We estimated the risk of experiencing SAEs in trials terminated for non-scientific reasons. This sub-analysis was exploratory in nature, and exclusion criteria had to be adapted during the course of the analysis. We included trials terminated for non-scientific reasons that reported summary results in ClinicalTrials.gov or EUCTR (German cohort). Summary results were only included if they provided data on SAEs in a tabular format. In both ClinicalTrials.gov and EUCTR, adverse event data are structured into separate sections for adverse events and SAEs. Despite some publications providing adverse event data, the inconsistency of reporting between registries and publications (25) prompted us to use only registry-based data for our analysis. We chose to focus on SAEs for this analysis because they are severe, clearly defined, and if summary results are reported, generally well-documented (9). To estimate the additional risk associated with the intervention, we compared the risk of SAEs in the intervention arm with the risk in the control arm. For this analysis, we included trials with at least two arms that could be clearly assigned to ‘control’ and ‘intervention’. Participants had to be randomized, and at least one participant had received an intervention and thus was at risk of experiencing an SAE. We excluded trials where intervention and control arms could not be assigned, such as single-arm trials, cross-over trials, or trials involving active comparators. We automatically extracted the number of patients at risk and the number of patients experiencing SAEs. We manually categorized the arms into control and intervention to calculate risk ratios and risk differences. For more details on the selection of eligible trials and the manual assignment of arms into control and intervention, see <u>appendix 4 (SAE risk analysis methodology).</u>

### Software

We used the cthist R package (17) to download the historical versions of trials from ClinicalTrials.gov. We developed the terminated-trial-analysis R package (26) to provide functions for this analysis. The R scripts and data generated for this study are available in the <u>terminated-trials-analysis-study repository</u>. Numbat Metanalysis Extraction Manager was utilized for data categorization in our study (27). The therapeutic focus of each clinical trial was assigned using the R TrialFociMapper package (19). Data cleaning steps and statistical analyses were performed using R [Version 4.3.2] (28).

## Results

### General trial characteristics

Out of 2,253 ClinicalTrials.gov registered trials in the German cohort, 1,670 (74%) were completed, and 217 (10%) were terminated. The remaining trials had other recruitment statuses (e.g., withdrawn, unknown) and were not included in the analysis. The 217 terminated trials had planned to enroll 37,379 participants but ultimately enrolled 14,723, resulting in a degree of enrollment of 39% at the time of termination. Among these terminated trials, 18 had enrolled more patients than planned. Completed trials had a degree of enrollment of 96%. Among terminated trials, 29% (n=64) reported summary results in ClinicalTrials.gov or EUCTR, 32% (n=69) as publications, and 35% (n=75) through either format. In comparison, 78% (n=1306) of completed trials reported results in any format. The median duration until termination was 1,026 days. In terms of therapeutic focus distribution among the terminated trials in the German cohort, neoplasms accounted for 14% of the distribution (n=60), followed by cardiovascular diseases at 10% (n=44), nervous system diseases at 7% (n=32), and infections at 2% (n=8). See table 1 (Characteristics of terminated trials and completed trials) for details.

Of the 1,091 trials in the Californian cohort with varied status, 921 (84%) were completed and 150 (14%) terminated. Trials with other recruitment statuses were excluded from the analysis. The enrollment percentage for terminated trials was 37%, compared to 83% for completed trials. The median duration until termination in the Californian cohort was 822 days. Among terminated trials, 62% (n=93) reported summary results in ClinicalTrials.gov, 32% (n=48) as publications, and 72% (n=108) through either format. In comparison, 87% (n=798) of completed trials reported results in any format. In terms of therapeutic focus distribution among the terminated trials in this cohort, neoplasms accounted for 23% (n=72) of the distribution, nervous system diseases represented 6% (n=17), cardiovascular diseases with 4% (n=12), and infections with 4% (n=11). For a complete breakdown of all therapeutic foci, see appendix table 2 (<u>Detailed therapeutic foci table</u>).

**Table 2:**
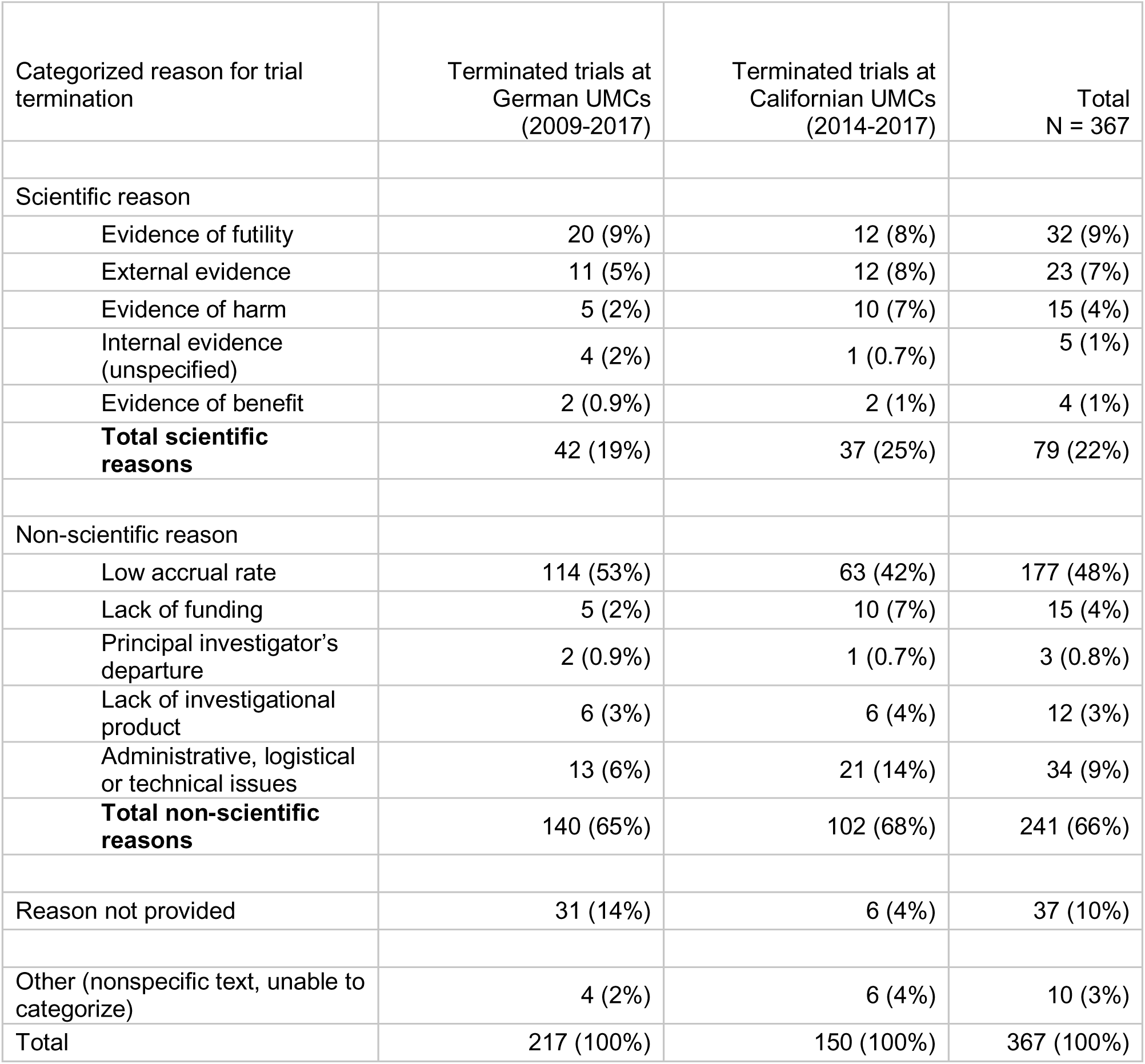
Reason for trial termination categorization.

| Categorized reason for trial termination | Terminated trials at German UMCs (2009-2017) | Terminated trials at Californian UMCs (2014-2017) | Total N = 367 |
| --- | --- | --- | --- |
| Scientific reason |  |  |  |
| Evidence of futility | 20 (9%) | 12 (8%) | 32 (9%) |
| External evidence | 11 (5%) | 12 (8%) | 23 (7%) |
| Evidence of harm | 5 (2%) | 10 (7%) | 15 (4%) |
| Internal evidence (unspecified) | 4 (2%) | 1 (0.7%) | 5 (1%) |
| Evidence of benefit | 2 (0.9%) | 2 (1%) | 4 (1%) |
| **Total scientific reasons** | **42 (19%)** | **37 (25%)** | **79 (22%)** |
| Non-scientific reason |  |  |  |
| Low accrual rate | 114 (53%) | 63 (42%) | 177 (48%) |
| Lack of funding | 5 (2%) | 10 (7%) | 15 (4%) |
| Principal investigator's departure | 2 (0.9%) | 1 (0.7%) | 3 (0.8%) |
| Lack of investigational product | 6 (3%) | 6 (4%) | 12 (3%) |
| Administrative, logistical or technical issues | 13 (6%) | 21 (14%) | 34 (9%) |
| **Total non-scientific reasons** | **140 (65%)** | **102 (68%)** | **241 (66%)** |
| Reason not provided | 31 (14%) | 6 (4%) | 37 (10%) |
| Other (nonspecific text, unable to categorize) | 4 (2%) | 6 (4%) | 10 (3%) |
| **Total** | **217 (100%)** | **150 (100%)** | **367 (100%)** |

### Reasons for trial termination

Among the 367 terminated trials across both datasets (Germany: 217, California: 150), 241 trials (66%) were terminated for non-scientific reasons, 79 trials (22%) for scientific reasons, and 37 trials (10%) did not provide a reason. Additionally, for 10 (3%) trials, incomplete or vague information made classification difficult (e.g., NCT01439958: ‘Core study 12011.201 was terminated’ and NCT01868503: ‘Protocol modification’). In these cases, raters were unable to determine the exact reason for termination and classified the trials into the “other” category. Overall, the reporting of reasons for trial termination was complete in the Californian cohort (96%) than in the German cohort (86%). See table 2 (Reason for trial termination categorization) for details.

For nine trials reporting both scientific and non-scientific reasons, the scientific reason was prioritized. The interrater reliability (Cohen’s kappa) was 83%. The most common nonscientific reason for termination was ‘low accrual rates’ (Germany: 53%,114; California: 42%,63), whereas the most frequently reported scientific reason was ‘evidence of futility’ (Germany: 9%, 20; California: 8%, 12).

In terms of enrollment, trials terminated for non-scientific reasons had an average enrollment of 34 participants (median: 14) and an average duration of 1092 days (median: 974), while those terminated for scientific reasons enrolled 127 participants (median: 25) with a duration of 1,001 days (median: 885). For further details, see appendix table 3 (<u>Trial characteristics by reason for termination</u>).

### SAE risk sub-analysis

Out of 241 trials terminated for non-scientific reasons, 76 (30%) had summary results reported in tabular format in EUCTR or ClinicalTrials.gov. Of these, 61 trials were excluded from the sub-analysis due to design or reporting limitations: 30 were single-arm trials, 20 had active comparators, 7 were non-randomized, 2 employed a crossover design, and 2 had not administered an intervention (see <u>PRISMA</u> <u>flow diagram 1</u>).

A total of 14 trials (ClinicalTrials.gov: 11, EUCTR: 3), in which a direct comparison of SAE risk between intervention and control arms was possible, were included in the sub-analysis. We found no statistically significant difference in experiencing SAE between patients in the control arm and patients in the intervention arm (see Figure 1). The common effect model showed a risk ratio (RR) of 1.0674 (95% CI: [0.8625; 1.3210], p = 0.5484), and the random effects model had an RR of 1.0459 (95% CI: [0.7599; 1.4396], p = 0.7830), with low-to-moderate heterogeneity (I² = 27.4%).

**Figure 1:**
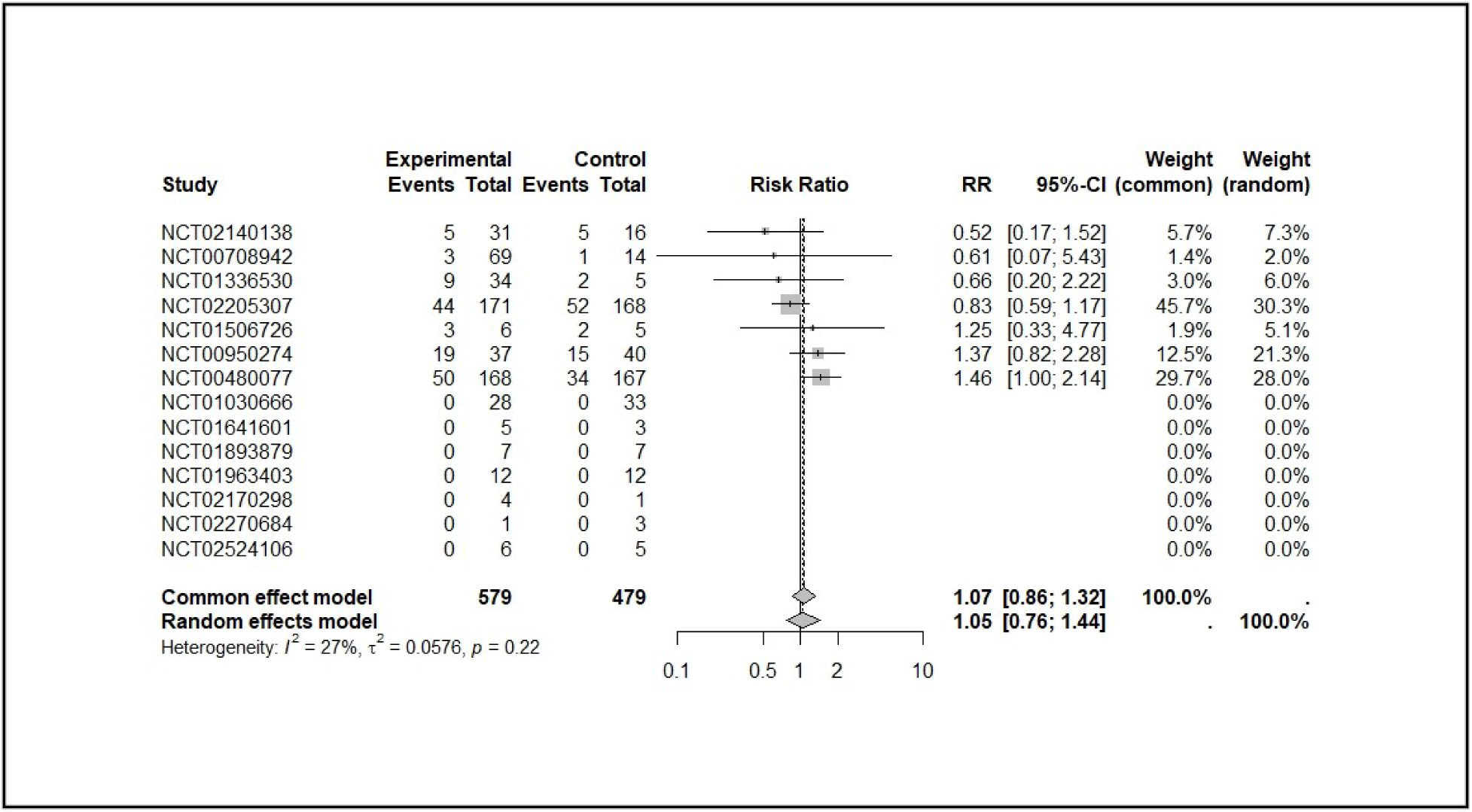
Comparison of serious adverse event risk between intervention and control groups in trials terminated for non-scientific reasons.

## Discussion

We developed scalable, semi-automated methods to investigate terminated trials registered in ClinicalTrials.gov, focusing on cohorts of clinical trials affiliated with German and Californian UMCs. Our analysis confirmed that terminated trials had lower rates of results reporting via any route (SR or publication) compared to completed trials, as seen in previous studies (1, 5). This gap reflects barriers to publishing results from terminated trials rather than limited registry reporting. Registries may serve as a valuable platform for disseminating findings from trials that are less likely to appear in journals. Across both datasets, over 60% of trials were terminated for non-scientific reasons, primarily due to low accrual rates. This aligns with previous studies emphasizing patient recruitment as the single main reason for early termination (5, 29). Overall, the reporting of reasons for trial termination in the free-text field of ClinicalTrials.gov was missing more frequently in the German cohort (31/217) compared to the Californian cohort (6/150).

Manual categorization of reasons for trial termination was relatively straightforward for the majority of trials and could potentially be automated. However, a subset posed challenges due to vague or insufficient termination descriptions. In these cases, the authors were unable to determine the exact reason for termination. Implementing predefined categories with an additional free-text option could enhance the analysis of trial terminations in registries. Registry data submitters and authors should also be encouraged to provide clearer explanations for termination, as this data can offer valuable insights for future trials. Since trial registries have limited space for specifying reasons, alternative ways to document key insights should be explored. Brageso et al. (2021) in publishing their findings on the terminated trial NCT03116165, highlighted lessons learned from their work, which were particularly relevant for future work in preventive psychological interventions for post-traumatic stress disorder in emergency settings (30).

We explored a methodology with the potential to be scaled up to assess the SAE risk using registry data for trials terminated due to non-scientific reasons. However, only 6% (14/242) of such trials could be included, as they reported results in the required tabular format on ClinicalTrials.gov or the EUCTR and had comparable intervention and control arms. Identifying eligible trials required extensive manual review, as automated categorization of trial arms was not feasible due to inconsistent labelling of trial arms. For instance, an identifier like ‘EG000’ in ClinicalTrials.gov is typically assumed to represent the experimental arm, but this assumption is incorrect in approximately 25% of cases, as noted in the description of AACT schema, which provides information on data elements in the registry (24). As a result, manual validation is required to ensure the accurate classification of trial arms.

The primary reason for the low inclusion rate was the limited availability of results reported in the structured tabular format required for our analysis (165/241). Many EUCTR records were excluded because results were submitted in non-tabular formats. While ClinicalTrials.gov mandates that sponsors of terminated trials with enrolled participants submit results including SAEs in a structured, tabular format, EUCTR permits the submission of PDF documents with varying content such as synopses, termination statements, statistical reports, or links. These inconsistencies hinder large-scale analyses and underscore the need for harmonized reporting standards.

Although the Consolidated Standards of Reporting Trials (CONSORT) guidelines (31) have improved reporting quality in academic publications, additional structured guidance is needed for clinical trial registries. A review by Chan et al. (2025) of 16 WHO primary registries including ClinicalTrials.gov and EUCTR revealed substantial variation in how summary results are presented (32). The new WHO guidance (2025) addresses this by outlining eight minimum elements, including results of benefits and harms, that should be reported in registries (33). Broader adoption of these standards could improve data consistency and facilitate the use of registry data for analytic purposes, including SAE assessments. Clearly defined reporting templates tailored to different study designs and interventions, together with strengthened quality control and internal logic checks, would further support both within-registry and cross-registry analyses, enhancing the feasibility of applying scalable methodological approaches for assessing SAE risks.

Our SAE analysis revealed no difference in the risk between the intervention and the control group. Previous studies have employed various classification approaches, such as grouping trials by intervention type or phase, which can lead to different results. For example, an analysis of lifestyle clinical trials (2000–2023) using data from ClinicalTrials.gov found no significant increase in the relative risk of experiencing an SAE in the intervention group compared to the control group (11). Another study examining phase 3 randomized controlled trials involving solid cancer patients comparing sorafenib with control found a significantly increased risk of SAEs in the intervention group compared to the control group (SAEs: RR 1.49, 95% CI: 1.18–1.89, p < 0.001) (30).

The methods we developed have their advantages. Our approach to characterize trials used historical versions of trial records to capture key dates, such as the primary completion date when trials were first marked as terminated. This approach provides insights into when termination decisions were initially recorded, offering a more updated timeline. Furthermore, our methods for assessing trial duration and enrollment percentage are scalable and can be applied to completed trials. Integrating these metrics with descriptive mappings of therapeutic areas and trial locations can contribute to exploratory analyses. While such data alone cannot determine trial feasibility or predict success, they may support preliminary assessments by identifying areas where early discontinuation has occurred and might indicate potential learnings from terminated trials.

While the methods were designed to maximize reproducibility and scalability, certain limitations, particularly those related to registry data quality and reporting inconsistencies, should be acknowledged. For the German and Californian cohorts, variables such as the availability of trial results publications were derived from the original datasets. As such, the possibility that some of these older trials have published results cannot be ruled out. While the assignment of reasons for trial termination and trial arms involved some degree of judgment, we implemented safeguards, including dual independent rating and resolution of disagreements via discussion. Nevertheless, some degree of misclassification cannot be ruled out. Therapeutic focus classification was performed using our TrialFociMapper tool, which retrieves and maps focus areas for trials registered in ClinicalTrials.gov. In some cases, a trial may be associated with multiple therapeutic foci. While users can manually review and annotate the most relevant focus, the tool does not yet perform automatic assignment of the most relevant therapeutic focus. In contrast, Brewster et al. (2022) manually reviewed study titles, abstracts, and descriptions for all included trials and assigned each to one of 29 predefined therapeutic foci (4). While our approach enables automated mapping, we acknowledge that assigning a single, most relevant focus remains an important step. Future development of the tool aims to incorporate an automated selection of the most relevant therapeutic focus per trial. Finally, for the SAE analysis, we relied solely on SAE data reported in trial registries. If SAE information was missing or incomplete in the registry, we did not use additional information from journal publications due to their non-standardized nature, with many studies reporting adverse events inconsistently between registries and publications (10, 34). We also did not search for additional information to determine whether SAEs were treatment-related.

In conclusion, our findings highlight that terminated trials report results less frequently than completed trials. Yet in a learning research system, data from ‘failed’ or terminated trials can provide valuable insights to inform future studies and help reduce inefficiencies in the clinical research system. Inconsistent reporting requirements hinder analysis, but broader adoption of harmonized standards across registries, structured templates, and registry logic checks could enable scalable assessments and enhance the transparency and utility of terminated trial data.

## Supporting information

### Data availability

All data used in analyses are publicly available at https://osf.io/n4ujs/ and https://github.com/sama9767/terminated-trials-analysis-study.

### Funding statement

Intramural funding was obtained for this study. The funders had no role in study design, data collection and analysis, decision to publish, or preparation of the manuscript

### Conflict of interest

No conflict of interest to declare.

### Author contributions

**SSY**: Conceptualization, Data curation, Formal analysis, Investigation, Methodology, Software, Visualization, Validation, Writing - original draft, Writing - review & editing. **BGC**: Conceptualization, Data curation, Formal analysis, Investigation, Methodology, Software, Supervision, Writing - review & editing. **DF**: Conceptualization, Methodology, Writing - review & editing. **MSH**: Methodology, Writing - review & editing. **DS**: Conceptualization, Methodology, Project Administration, Supervision, and Writing – Review & Editing. **SGS**: Conceptualization, Data curation, Formal analysis, Investigation, Methodology, Project Administration, Supervision, Validation, Visualization, Writing – Review & Editing.

## Supporting information

Protocol

STROBE checklist

## Notes

### Competing Interest Statement

The authors have declared no competing interest.

### Clinical Protocols

https://osf.io/n4ujs

### Summary of Updates

This version includes a correction to the author name, which was mistakenly listed incorrectly in the previous version.

