## Supplementary file for "Exploring scalable assessment methods for terminated trials in ClinicalTrials.gov: A cohort analysis of German and Californian trials"

Supplementary material

### Appendix 1: Degree of enrolment - Handling of missing enrollment variable

The enrollment variable in ClinicalTrials.gov differentiates between anticipated (estimated) and actual enrollment based on the reported enrollment type. In cases where the enrollment function flagged a "missing data" warning, the trial versions were manually reviewed and handled in the following way:

Example 1: The enrollment type was not defined, but the enrollment number suggests an anticipated value. These cases were manually reviewed, the anticipated enrollment was inferred, and the data were included in the analysis.

Trial: NCT00150878

| Historical version | Enrollment type | Enrollment |
| --- | --- | --- |
| 1 | NA | 172 |
| 2 | NA | 172 |
| 3 | NA | 172 |
| 4 | NA | 172 |
| 5 | actual | 198 |
| 6 | actual | 198 |

Example 2: Enrollment type was recorded as “actual” across all versions, and anticipated enrollment could not be determined. These cases were excluded from the analysis.

Trial: NCT01030666

| Historical version | Enrollment type | Enrollment |
| --- | --- | --- |
| 1 | actual | 61 |
| 2 | actual | 61 |
| 3 | actual | 61 |
| 4 | actual | 61 |
| 5 | actual | 61 |
| 6 | actual | 61 |
| 7 | actual | 61 |
| 8 | actual | 61 |

Example 3: Both enrollment type and enrollment numbers are missing (NA). These incomplete cases were excluded from the analysis.

Trial: NCT01071135

| Historical version | Enrollment type | Enrollment |
| --- | --- | --- |
| 1 | NA | NA |
| 2 | actual | 5 |

### Appendix Table 2. Reason for termination - categorization

|  | Categorized reason for termination | Explanation |
| --- | --- | --- |
| **1** | **Scientific reason for trial termination** | |
| 1a | Evidence of harm  (other terms: safety, toxicity, harm) | The risk for participants unexpectedly outweighs the benefits. Reasons include severe adverse events and/or the negative impact of the intervention based on an interim analysis. |
| 1b | Evidence of benefit  (other terms: benefit) | The new treatment shows a clinically highly significant beneficial effect in the interim analysis. |
| 1c | Evidence of futility  (other terms: futility, relevance loss) | The study hypothesis is unexpectedly shown to be unprovable within the constraints of the trial based on an interim analysis. |
| 1d | External evidence  (other terms: SOC change, external evidence) | Changes in the standard of care, the disease/condition no longer exists, or results from other trials lead to trial termination. |
| 1e | Internal evidence (unspecified)  (other terms: sufficient (safety) information collected,  recommendation by a regulatory body after interim analysis) | The study has terminated for scientific reasons, but there is insufficient information to determine whether it would fall into 1b or 1c. |
| **2** | **Non-scientific reason for trial termination** | |
| 2a | Low accrual rate  (other terms: slow/poor/low enrollment, dropout, recruitment issue, accrual failure) | Lack of participants enrolling in the trial. |
| 2b | Lack of funding  (other terms: external/internal funding, funding not secured, expensive, sponsor back out) | Insufficient external or internal funding, funding not secured, or the sponsor withdrawing support. |
| 2c | Principal investigator’s departure  (other terms: PI left, investigator left) | The departure of the principal investigator from the trial. |
| 2d | Lack of investigational product  (other terms: manufacturing issue, product no longer available, product expired/contaminated) | Issues related to manufacturing, including the unavailability, expiration, or contamination of the investigational product (drug or device). |
| 2e | Administrative, logistical or technical issues  (other terms: administrative issue, logistic failure, technical failure, changes in study planning, sponsor/company decision) | Challenges related to management, operational processes, or technical aspects of the study other than those non-scientific reasons specified above (2a,2b,2c,2d). |
| **3** | **Reason not provided** | No information was given by data submitters. |
| **4** | **Other** | Information available but unspecific or uninformative includes trials that were completed normally but were marked as terminated for administrative reasons or in error. |

### Appendix Table 3. Detailed therapeutic foci table

| **Therapeutic foci ^1^** | Completed trials at Californian UMCs  (2014-2017) | Completed trials at German UMCs  (2009-2017) | Terminated trials at Californian UMCs  (2014-2017) | Terminated trials at  German UMCs  (2009-2017) |
| --- | --- | --- | --- | --- |
| Behavior and Behavior Mechanisms | 41 (2%) | 51 (2%) | 2 (1%) | 2 (0%) |
| Cardiovascular Diseases | 80 (5%) | 280 (10%) | 12 (4%) | 44 (10%) |
| Chemically Induced Disorders | 36 (2%) | 9 (0%) | 3 (1%) | 2 (0%) |
| Congenital, Hereditary, and Neonatal Diseases and Abnormalities | 53 (3%) | 69 (2%) | 5 (2%) | 7 (2%) |
| Digestive System Diseases | 51 (3%) | 113 (4%) | 14 (5%) | 28 (6%) |
| Endocrine System Diseases | 53 (3%) | 73 (2%) | 2 (1%) | 14 (3%) |
| Eye Diseases | 16 (1%) | 64 (2%) | 4 (1%) | 2 (0%) |
| Hemic and Lymphatic Diseases | 37 (2%) | 94 (3%) | 23 (7%) | 18 (4%) |
| Immune System Diseases | 82 (5%) | 132 (4%) | 18 (6%) | 23 (5%) |
| Infections | 66 (4%) | 101 (3%) | 11 (4%) | 8 (2%) |
| Mental Disorders | 138 (8%) | 149 (5%) | 7 (2%) | 9 (2%) |
| Musculoskeletal Diseases | 27 (2%) | 49 (2%) | 4 (1%) | 5 (1%) |
| Neoplasms | 160 (9%) | 261 (9%) | 72 (23%) | 60 (14%) |
| Nervous System Diseases | 132 (8%) | 222 (8%) | 17 (6%) | 32 (7%) |
| Nutritional and Metabolic Diseases | 105 (6%) | 109 (4%) | 6 (2%) | 15 (3%) |
| Occupational Diseases | 1 (0%) | 2 (0%) | 0 (0%) | 0 (0%) |
| Otorhinolaryngologic Diseases | 10 (1%) | 39 (1%) | 0 (0%) | 2 (0%) |
| Pathological Conditions, Signs and Symptoms | 233 (13%) | 416 (14%) | 38 (12%) | 67 (15%) |
| Respiratory Tract Diseases | 54 (3%) | 102 (3%) | 13 (4%) | 21 (5%) |
| Skin and Connective Tissue Diseases | 63 (4%) | 110 (4%) | 23 (7%) | 23 (5%) |
| Stomatognathic Diseases | 11 (1%) | 32 (1%) | 2 (1%) | 2 (0%) |
| Urogenital Diseases | 100 (6%) | 88 (3%) | 16 (5%) | 18 (4%) |
| Wounds and Injuries | 26 (1%) | 40 (1%) | 3 (1%) | 5 (1%) |
| Missing Foci Entry | 161 (9%) | 340 (12%) | 13 (4%) | 29 (7%) |

**^1^**Trials can be assigned to multiple therapeutic categories; percentages reflect the proportion of total occurrences rather than unique trials.

### Appendix Table 4. Trial characteristics by reason for termination

| Reason for termination | No. of Trials | Average Enrollment | Median Enrollment | Avg. Duration (Days) | Median Duration (Days) |
| --- | --- | --- | --- | --- | --- |
| Scientific reason | 79 (21%) | 127 | 25 | 1001 | 885 |
| Non-scientific reason | 241 (66%) | 34 | 14 | 1092 | 974 |
| Reason not provided | 37 (10%) | 98 | 30 | 1051 | 915 |
| Other | 10 (3%) | 155 | 17 | 738 | 518 |
| **Total** | 367 (100%) | - | - | - | - |

### Appendix 5. SAE risk analysis methodology

**Trial selection and data extraction**

In the SAE analysis, we included trials terminated for non-scientific reasons, having summary results reported in a tabular format. For trials lacking results in Clinicaltrials.gov and being cross-registered in the EU Clinical Trials Register (EUCTR), we used a custom web scraper to check for the availability of summary results. For ClinicalTrials.gov trials that reported results in the registry, we used ‘get_ctgov_sae_group_data’ function from [terminated-trials-study](https://github.com/sama9767/terminated-trials-study) R package to extract the **number of patients at risk** (‘At risk’) and the **number of patients experiencing serious adverse events** (‘Affected’) for all arms of the trials, including their descriptions and group IDs.

For EUCTR trials, we manually included those with results available in a tabular format. Trials where results were provided only via external links, such as publications, final statements, synopses, statistical reports, or multiple links, were excluded. For included EUCTR trials, we used a scraper function from [terminated-trials-study](https://github.com/sama9767/terminated-trials-study) R package to automatically extract from the 'Adverse events' table the **number of subjects affected by serious adverse events** ('Total subjects affected by serious adverse events') and **the number of subjects exposed** ('Total subjects exposed by serious adverse events').

**Inclusion and exclusion criteria**

We included trials that met the following criteria:

- Had at least two arms where both intervention and control groups could be assigned.
- Randomized participants (this criterion was added during the process).
- Included at least one participant received an intervention and was at risk of experiencing SAE (this criterion was added during the process).

We excluded trials if:

- They included only one treatment arm.
- They used active comparators rather than a control group.
- They were cross-over trials where participants received both the control and intervention treatments in a randomized order (this criterion was added during the process).

**Manual categorization of arms**

Two authors (SGS and SSY) independently assigned intervention and control arms. Disagreements were resolved through discussion or by consulting a third author (BGC). The classification was guided by the following criteria:

- Control Arm: Included placebo, sham, or no-intervention groups.
- Intervention Arm: Included experimental or access groups.

Using the information collected in the previous steps, we combined multiple intervention and control arms into a single intervention and control group where applicable. This combined data was then used to calculate the risk ratio and risk difference for serious adverse events between the intervention and control groups. Following is the PRISMA flow diagram to illustrate the selection procedure.

### PRISMA flow diagram 1

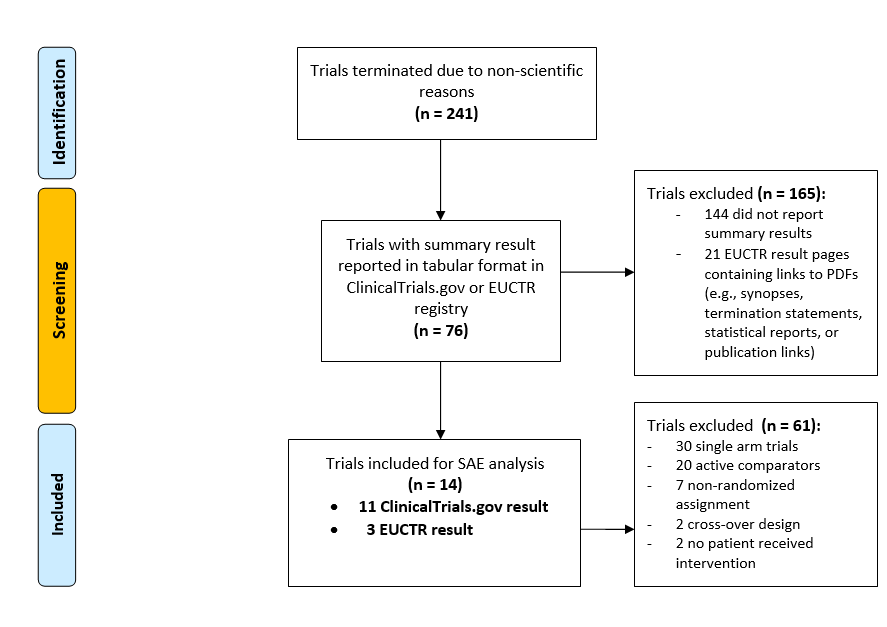
