## Supplementary material for "Exploring scalable assessment methods for terminated trials in ClinicalTrials.gov: A cohort analysis of German and Californian trials": Protocol

Samruddhi Yerunkar (SSY)[1], Susanne Gabriele Schorr (SGS)[1], Benjamin Gregory Carlisle (BGC)[2], Delwen Franzen (DF)[1,] Maia Salholz-Hillel (MSH)[1], Daniel Strech (DS)[1]

1. QUEST Center for Responsible Research, Berlin Institute of Health, Germany

2. STREAM research group, Department of Ethics, Equity and Policy, School of Population and Global Health, McGill University, Montréal, Canada

### Background

Clinical trials are the cornerstone of evidence-based healthcare, providing essential information on the safety and efficacy of new treatments and interventions. However, not all trials reach their pre-defined intended goals and many discontinue prematurely or terminate. Speich et al., (2022) analyzed 326 trials approved by research ethics committees in Switzerland, the United Kingdom (UK), Germany, and Canada, and reported that 30% (98/326) of trials were prematurely discontinued. The study also highlights that such trials were less likely to have their results published in journals or registries compared to completed trials (1), thereby preventing participants' efforts from contributing to scientific knowledge.

Premature discontinuation of trials can occur due to scientific reasons such as ethical concerns related to efficacy or safety, or non-scientific reasons such as poor project planning or low enrollment rates (2, 3). Williams et al., (2015) highlighted that, as of February 2013, the most common cause of trial termination among terminated trials posted on ClinicalTrials.gov was non-scientific factors (3). This is particularly concerning because these issues could in principle have been foreseen or prevented, leading to an unnecessary expenditure of time and resources. Most importantly, participants who invest their valuable time to participate in clinical research might be exposed to interventions and measures with unclear benefits, and risk experiencing adverse events.

Clinical trial interventions are often experimental and carry a higher risk of adverse events than standard clinical practice (4). Specifically, serious adverse events (SAEs) are defined as “any untoward medical occurrence that at any dose results in death, is life-threatening, requires inpatient hospitalisation or prolongation of existing hospitalisation, results in persistent or significant disability/incapacity, or is a congenital anomaly/birth defect” (5), and must be always fully reported (6, 7). Studies have consistently shown that reporting of these SAEs is significantly more comprehensive on ClinicalTrials.gov compared to published journal articles (7-9). By analyzing the SAEs data from ClincialTrials.gov, and comparing intervention to control group, researchers can gain valuable insights into risks participants experienced in trials. .

Our group recently launched a dashboard presenting how clinical trials conducted at University Medical Centers (UMCs) in Germany perform on several responsible research practices, such as prospective registration and results reporting (10). The dashboard also shows the breakdown of these practices by UMCs, allowing them and other interested parties to identify and monitor those areas of responsible research where improvement is necessary. In its current form, the dashboard does not provide information on how many trials were prematurely discontinued. In addition to the German UMCs dashboard, our group has conducted an ongoing study in six Califonia medical schools, assessing transparency practices in the US and it also does not include information on prematurely discontinued trials (11).

This study aims to identify efficient and scalable methods to characterise terminated trials. We will describe the characteristics of terminated trials, compare, and quantify reasons for trial termination in the two samples of clinical trials conducted at UMCs in Germany and in California (US). In a subsample of trials terminated due to non-scientific reasons, we aim to explore measures to estimate patient harm by comparing the risk of experiencing a serious adverse event for a patient in the intervention group with the control group.

### Methods

#### Data sources and sample

The study will use two datasets:

1) German UMCs: Two related projects (IntoValue and IntoValue2), have drawn a sample of registry entries for interventional studies conducted by German UMCs and reported as complete between 2009 and 2017 (12-14). The trials were registered in ClinicalTrials.gov or the German clinical trials register, Deutsches Register Klinischer Studien (DRKS). The registry data was downloaded on 1 November 2022. We will retrieve the combined dataset from GitHub: https://github.com/maia-sh/intovalue-data, where the IntoValue dataset is actively maintained. Only trials registered in ClinicalTrials.gov will be included.

2) Californian UMCs (CONTRAST): Interventional clinical trials registered on ClinicalTrials.gov conducted at California medical schools and reported as complete between 2014 and 2017 (15).

#### Eligibility criteria

We will only focus on trials registered in ClinicalTrials.gov with the recruitment status 'Terminated' at the registry download date. ClinicalTrials.gov defines 'terminated' as trials where the recruitment or enrollment of participants has halted prematurely and will not resume, and participants are no longer being examined or receiving intervention. Additionally, we only included trials with a minimum enrollment of at least one participant in our sample.

#### Data extraction and analyses

##### *Characteristics of terminated trials*

We will obtain trial identifiers (NCT number) and other basic information (phase, study type, etc.) about terminated trials from the two datasets. We will give a baseline description of the two samples of terminated trials. Additionally, we will link the manual searched information on results reporting from the IntoValue and the CONTRAST dataset to describe the number of terminated trials with publications.

To download the historical clinical trial registry entry data, we will use the cthist R package (16), which allows the mass-downloading of such data. By analyzing the historical changes throughout the trial lifecycle, we will generate the variables ‘degree of enrollment’ and ‘duration of trial’. During the lifecycle of the trial, the enrollment variable begins as “estimated” at the start of the trial and it is later updated to “actual” at various stages of the trial, including its completion. We consider anticipated enrollment as estimated enrolment reported in the first version on or after the trial's start date launch and actual enrollment as final enrollment recorded at the end of the trial. The ‘degree of enrollment’ variable is defined by the ratio between actual and anticipated enrollment, expressed as a percentage of enrollment achieved until termination.

The variable ‘trial days’ describes the number of days between the ‘stop date’ (the actual primary completion date reported on the clinical trial record where its overall status was first changed to ‘Terminated’ from any other overall status in the ClinicalTrials.gov registry) and the ‘start date’ (start date taken from the same version of the trial record when the trial was updated as ‘Terminated’ from any other status in the ClinicalTrials.gov registry).

***Summary results reporting***

For both datasets, we will report if summary results are available on ClinicalTrials.gov. For trials within the German dataset that indicate cross-registration in the EUCTR registry and lack summary results in ClinicalTrial.gov, we will manually check if summary results are available in EUCTR. Summary results reporting in this sample will be considered as true if summary results are present in either ClinicalTrials.gov or the EUCTR registry.

All code to generate variables characterizing trials discontinued prematurely will be made available on https://github.com/sama9767/terminated-trials-study.

***Reason for termination***

The "why_stopped" field extracted from Clinicaltrials.gov using the cthist package provides a brief explanation of the reasons for study termination. It is a free-text field limited to 160 characters.

Based on published literature and web resources (1-3, 17-21), we developed and iteratively refined a categorization table for reasons for termination:

(1) Termination based on scientific data collected during the trial (scientific): Termination due to reasons like findings related to the overall benefit-risk profile of intervention or external evidence.

(2) Termination based on a reason other than scientific data (non-scientific): This category encapsulates potentially foreseeable or preventable reasons like poor recruitment or organizational failure.

(3) Reason for termination not provided (unknown): If the reason for termination is not provided in the registry.

Table 1: Reason for termination classification

| Termination category | Description |
|---|---|
| **1. Termination based on scientific data collected** | |

| | |
|---|---|
| 1a. Evidence of harm | The risk for participants unexpectedly outweighs the benefits. Reasons include severe adverse events, and/or the negative impact of the intervention based on an interim analysis<br>Other terms: safety, harm, toxicity |
| 1b. Evidence of benefit | The new treatment shows a clinically highly significant beneficial effect in an interim analysis.<br>Other terms: benefit |
| 1c. Evidence of futility | The study hypothesis is unexpectedly shown to be unprovable within the constraints of the trial based on an interim analysis.<br>Other terms: futility |
| 1d. External evidence | Changes in standard of care, disease/condition no longer exist, results from other trials<br>Other terms: Change SOC, external evidence |
| **2. Termination based on a reason other than scientific data** | |
| 2a. Low accrual rate | Poor/slow enrollment<br>Other terms: lack of participation, dropout, recruitment, accrual |
| 2b. Lack of funding | Lack of funding<br>Other terms: funding not secured, external/internal funding, expensive |
| 2c. Principal Investigator's departure | Principal Investigator leaving the trial<br>Other terms: PI left, investigator left |
| 2d. Lack of investigational product | Manufacturing or other issues related to investigational product.<br>Other terms:<br>product discontinued, product withdrawal |
| 2e. Administrative, logistical or technical issues. | Challenges related to management, operational processes, or technical aspects of the study other than those specified above (1a.-1d; 2a-2d)<br>Examples include changes in study planning, technical failure/unresolved issues contributing to trial termination, sponsor/company decision |

| | |
|---|---|
| 2f. Other (e.g., nonspecific text, unable to categorize) | Information available, but unspecific or uninformative |
| **3. Termination reason not provided (unknown)** | |
| Unknown | No information available |

Two authors (SGS and SSY) will categorize the reason for termination independently of each other using Numbat Metanalysis Extraction Manager (22)**.** All reasons will be documented. The primary reason will be identified using the following procedure: If non-scientific and scientific reasons are mentioned, the scientific reason will be considered primary. If multiple reasons within the same category are mentioned, the primary reason will be defined by the wording used to describe the reason and if this is not sufficiently clear the sequence will be used as a reference. Discrepancies between authors will be solved by discussion, and if no agreement is reached a third person will be consulted. The categorization has been tested in a small pilot categorizing a random sample of 20 trials per dataset. Minor updates were conducted based on the results of the pilot.

***Subsample analysis***

In the subsample of trials terminated due to non-scientific reasons, we aim to estimate the risk of experiencing a serious adverse event for patients in the intervention group compared to patients in the control group. To achieve this, we will include all trials reporting on adverse events in ClinicalTrials.gov that have at least two arms (and where the combination of different intervention arms is possible). Trials with an active comparator will be excluded. For all trials, we will automatically extract the number of patients at risk and the number of patients experiencing serious adverse events and manually link this data to the relevant arm.

All adverse event information collected from the intervention and control groups will be utilized to calculate risk ratio and risk difference. We will then meta-analyse this using the **metabin()** function from the R programming language's "meta" package (23). This approach will allow us to estimate the additional risk for patients in the intervention group compared to the control group, highlighting the patient harm in trials that were later discontinued due to potentially foreseeable or preventable reasons like poor recruitment or organizational failure.

**Software**

We will use the R package aactr (24) to download information from the AACT database (25) and cthist (16) to download the historical versions of trials. We will use web scrapers to access information from the EUCTR registry (26-28). Numbat Metanalysis Extraction Manager will be utilized for data categorization in our study (22). Data cleaning steps and statistical analyses will be performed using R (29).

Exploring scalable assessment methods for terminated trials in ClinicalTrials.gov: Cohort analysis in Germany and California

### Code Availability

All analysis scripts will be made openly available from https://github.com/sama9767/terminated-trials-analysis-study.

### Supplementary information

For each dataset, we will present trial-level information. The provided information will include trial id (nct id), center_size, main_sponsor, is_prospective, masking, phase, therapeutic foci, reason for termination, summary results availability, start date, stop date, trial days (n), anticipated enrollment, actual enrollment, degree of enrollment (%), and the number of patients at risk for and affected by SAE in the intervention and control group (wherever reported).
